# Unravelling the Complex Inflammatory Landscape of COVID-19 infection: A Pathway to Biomarkers Identification in Infection-Associated Delirium in the ICU

**DOI:** 10.1101/2025.03.06.25323473

**Authors:** Tristan Born, Matthieu Perreau, Pierre-Paul Axisa, Craig Fenwick, Andrea Pinto, Nawfel Ben-Hamouda, Andrea O. Rossetti, Renaud Du Pasquier, Jean-Daniel Chiche, Raphaël Bernard-Valnet

**Affiliations:** Service of Neurology, Department of Clinical Neurosciences, Lausanne University Hospital (Centre Hospitalier Universitaire Vaudois) and University of Lausanne, Switzerland; Service of Immunology and Allergy, Department of Medicine, Lausanne University Hospital (Centre Hospitalier Universitaire Vaudois) and University of Lausanne, Switzerland; Cancer Research Center of Toulouse (CRCT), Institut National de la Santé et de la Recherche Médicale (INSERM), Centre National de la Recherche Scientifique (CNRS), Université Toulouse III-Paul Sabatier (UPS), Toulouse, France; Department of Computer Science (INFK), ETH Zurich, Zurich, Switzerland; Department of Intensive Care Medicine, Lausanne University Hospital (Centre Hospitalier Universitaire Vaudois) and University of Lausanne, Switzerland

## Abstract

**Background:** Delirium is a serious complication in patients with COVID-19-related acute respiratory distress syndrome (ARDS) admitted to the intensive care unit (ICU). Although numerous clinical risk factors have been identified, the immunologic pathways underlying delirium remain unclear. In this retrospective cohort study, we investigated high-dimensional immune signatures in ICU patients to delineate peripheral immune markers associated with delirium. We also explored machine learning (ML) approaches to enhance biomarker discovery and strengthen predictive modelling through synthetic data generation.

**Methods:** Sixty-two COVID-19 ARDS patients admitted to the ICU at Lausanne University Hospital, Switzerland, were studied; 39 (63%) developed delirium. Control cohorts consisted of 55 non-ICU COVID-19 patients and 450 healthy individuals. We performed high-dimensional immunophenotyping of cytokines, chemokines, and growth factors using multiplex beads assay, along with immune cell profiling via mass cytometry (CyTOF). Ridge regression has been employed to build classification models. To address the limited sample size and improve model stability, we generated a synthetic dataset using beta-variational autoencoders.

**Results:** Delirious patients exhibited a distinctive immune signature, including elevated CXCL1, CCL11, CXCL13, HGF, and VEGF-A, coupled with reduced IL-1α, IL-21, and IL-22. Alterations in immune cell populations featured increased exhausted B cells and decreases in CXCR3+ CD4+ T cells, IgM+ unswitched memory B cells, and HLA-DR+ activated T cells. Leveraging these high-dimensional data, we trained ridge regression models to predict delirium. Incorporating synthetic data helped stabilize the models with a best-performing model achieving an area under the curve (AUC) of 0.95, with high sensitivity (93%) and specificity (86%), based on 12 identified markers.

**Conclusions:** Our findings demonstrate a distinct immune profile linked to ICU delirium and illustrate how ML can enhance biomarker discovery. Further prospective validation may refine these markers and guide precision-targeted interventions for mitigating delirium in critically ill populations.

**Graphical Abstract**

## Introduction

Delirium is a common neuropsychiatric syndrome in hospitalized patients, particularly in intensive care unit (ICU), where its prevalence can reach 50-70% in mechanically ventilated patients [1]. It is characterized by altered cognition, disorganized thinking, and fluctuating arousal. Delirium is linked with poor outcomes, such as prolonged hospital stays, increased mortality, and cognitive decline [2, 3] related to neuronal damage, as indicated by elevated serum neurofilament light chain (NfL) levels [4]. Various predisposing and precipitating factors contribute to delirium, especially in the ICU, where environmental and host-related factors, such as mechanical ventilation, pharmacological sedation, and metabolic disturbances, play a role. Neurobiological mechanisms involve systemic and neuroinflammation, neurotransmitter imbalances, and vascular dysfunction [1].

The role of pro-inflammatory cytokines, such as IL-6 and TNF-α, in the pathophysiology of delirium has been highlighted, but the exact pathways involved remain unclear [1]. SARS-CoV-2 has exacerbated infection-related delirium, with up to 80% of ICU COVID-19 patients affected [5, 6]. Although COVID-19 is primarily respiratory, delirium is likely driven by an immune-mediated response rather than direct CNS invasion [7]. Hyper-inflammatory states, characterized by elevated IL-1β, IL-6, TNF, and IL-8 levels, may disrupt the blood-brain barrier (BBB), leading to neuroinflammation and neuronal injury [8–11]. However, the precise immune signature responsible for delirium is still debated, and no prognostic biomarkers have been implemented into clinic [12].

Although currently recommended tools such as the Confusion Assessment Method for the Intensive Care Unit (CAM-ICU) [13] and the Intensive Care Delirium Screening Checklist (ICDSC) [14] allow for delirium assessment, pharmacologic options remain somewhat limited [15], while preventive non-pharmacological interventions can reduce the incidence of delirium by up to 40% [16].

Our study aimed to investigate peripheral immune mediators (cytokines, chemokines, and growth factors) and leukocytes contributing to delirium in the ICU, using COVID-19 as a representative disease of infection-associated delirium. We also sought to determine whether a combination of these inflammatory markers could help predict patients at risk to develop delirium.

## Methods

### Design, Setting, and Participants

We utilized data generated from two studies previously conducted at Lausanne University Hospital (Centre Hospitalier Universitaire Vaudois [CHUV]) to perform this work. The first study provided clinical data on ICU stays and delirium, extracted from a cohort of 311 consecutive adult patients (≥18 years old) hospitalized in the ICU between December 2017 and June 2021, all of whom met the Berlin criteria for ARDS (253 with COVID-19 and 58 with other causes). Delirium in these patients was assessed using the CAM-ICU, routinely performed by nurses twice a day in patients with a RASS score of ≥-2. Patients were considered delirious if they had at least one positive CAM-ICU assessment during ICU stay. The data collection process has been detailed previously [5].

From the 231 patients with CAM-ICU assessments (**Supp. Fig. 1**), we included all COVID-19 patients who had assessments of soluble inflammatory markers by Luminex and immunophenotyping by mass cytometry from one of our previous studies, which aimed to define the key serum immune signatures associated with COVID-19 severity [17].

As controls, we included 55 COVID-19 patients who did not require ICU hospitalization and 450 healthy individuals.

### Assessment of serum immune profile and Immunophenotyping

Concentration of serum soluble factors (cytokines, chemokines, growth factors) was performed by multiplex bead assay and in depth immunophenotyping by mass cytometry (CyTOF) were performed as described previously [17]. Correspondence between cell populations and assessed surface markers is provided in **Supp. Table 1**. A detailed description is provided in the **Supplementary Appendix**.

### Assessment of other variables

Data related to the ICU stay were extracted directly from patient’s electronic medical records (MetaVision, iMDSoft, Tel Aviv, Israel). Data collected included demographics (age and sex), weight, body mass index (BMI), Simplified Acute Physiology Score II (SAPS II) on admission, PaO_2_/FiO_2_ ratio on admission, worst PaO_2_/FiO_2_ ratio, confusion assessment method (CAM-ICU) (which was routinely assessed twice a day only during the ICU stay), total dose (including boluses and continuous infusions) of fentanyl, propofol, and midazolam, coma duration (defined as the number of days with a Richmond Agitation–Sedation Score [RASS] at −4 or −5), duration of the say in the ICU, duration of mechanical ventilation, steroid, tocilizumab, and hydroxychloroquine use.

### Handling of missing values

Patients with missing data were excluded from this study and no data were to be imputed.

### Statistical Analysis

According to STROBE (Strengthening the Reporting of Observational Studies in Epidemiology) guidelines, continuous variables were reported as median (interquartile range [IQR]: 25%–75%) and categorical variables as numbers and percentages. Comparisons across groups were performed using the chi-squared test for categorical variables, and the Student’s t-test or the Mann-Whitney U-test in continuous variables, as appropriate. Correlation coefficients were calculated using the Spearman rank coefficient due to the violation of normality assumptions and the presence of numerous outliers. All p-values reported are two-sided, and statistical significance was set at α = 0.05. A detailed description of all statistical approaches and R packages used is provided in the **Supplementary Appendix**.

### Standard Protocol Approvals, Registrations, and Patient Consents

This study has been approved by the local ethic committee (CER-VD) in the frame of the CORO-NEURO study (authorization no. 2020-01123). We obtained a consent waiver for deceased patients.

### Data availability

The grouped high-dimensional analyses and the code used for figure generation and modelling are available at https://github.com/rbernardvalnet/delirium_prediction.git. The anonymized clinical data that support the findings of this study can be obtained from the authors upon reasonable request by qualified, noncommercial researchers. The data are not publicly available due to privacy restrictions.

## Results

### Clinical presentation and Delirium in ICU-admitted patients

Overall, 62 patients admitted to the ICU for ARDS associated with COVID-19 were included in the final analysis (**Supp. Fig. 1**). Among them, 39 (63%) developed delirium (positive CAM-ICU) during their stay, with a median duration of 3 days (IQR 2-5) (**Table 1**). The baseline characteristics of these patients were generally similar between those who developed delirium and those who did not, in terms of age, sex, BMI, and severity at admission (SAPSII score).

**Table 1.** Demographic and clinical characteristics of COVID19 patients hospitalized in the intensive care unit.

| Variable | Delirium (n = 39) | No delirium (n = 23) | p-value |
| --- | --- | --- | --- |
| **Baseline characteristics** |  |  |  |
| Age in year, median (IQR) | 60 (56-65) | 66 (60-74) | 0.068 |
| Sex, female, n (%) | 7 (18%) | 10 (43%) | 0.060 |
| BMI kg.m<sup>-2</sup>, median (IQR) | 28 (26-32) | 27 (25-30) | 0.198 |
| **ICU admission** |  |  |  |
| SAPS II at admission, median (IQR) | 37 (31-46) | 36 (33-41) | 0.323 |
| **ICU course and outcome** |  |  |  |
| Coma<sup>a</sup>, number of days, median (IQR) | 12 (9-20) | 2 (0-7) | < **0.001** |
| Stay in the ICU, number of days, median (IQR) | 20 (16-36) | 9 (5-12) | < **0.001** |
| Positive CAM-ICU, number of days, median (IQR) | 3 (2-5) | 0 | < **0.001** |
| **Respiratory support** |  |  |  |
| Worst PaO<sub>2</sub>/FiO<sub>2</sub>, median (IQR) | 0.57 (0.52-0.63) | 0.64 (0.58-0.81) | **0.015** |
| Time of mechanical ventilation in days, median (IQR) | 15 (11-27) | 5 (0-11) | < **0.001** |
| **Analgo-sedative drugs, total dose received during ICU stay** |  |  |  |
| Total midazolam doses in mg/kg, median (IQR) | 3.82 (0.08-13.09) | 0 (0-0.56) | **0.001** |
| Total propofol doses in mg/kg, median (IQR) | 917 (496-1339) | 246 (0-562) | < **0.001** |
| Total fentanyl doses in ug/kg, median (IQR) | 343 (211-560) | 46 (0-177) | < **0.001** |
| **Antiviral and anti-inflammatory treatment** |  |  |  |
| Received corticosteroids<sup>b</sup> before sampling, n (%) | 18 (46%) | 8 (35%) | 0.541 |
| Received tocilizumab before sampling, n (%) | 24 (62%) | 14 (61%) | 0.939 |
| Received hydroxychloroquine before sampling, n (%) | 19 (48%) | 12 (52%) | 0.792 |
| **Multimodal inflammatory testing** |  |  |  |
| Time between ICU admission and sampling, median (IQR) | 2 (1-6) | 2 (1-8) | 0.994 |

Abbreviation: IQR = interquartile range; BMI = body mass index; CAM-ICU: Confusion Assessment Method for the Intensive Care Unit; ICU = intensive care unit; SAPS II = Simplified Acute Physiology Score II; FiO<sub>2</sub> = inspired oxygen fraction; PaO<sub>2</sub> = Arterial partial pressure of oxygen.
<sup>a</sup>: Defined as the number of days with a Richmond Agitation–Sedation Score [RASS] at -4 or -5.
<sup>b</sup>: Dexamethasone, prednisolone, or methylprednisolone.

As expected, delirium was more frequent in patients with worse respiratory support parameters, including a longer duration under mechanical ventilation (median [IQR]: 15 [11–27] vs. 5 [0–11] days, p < 0.001) and a lower PaO_2_/FiO_2_ ratio (median [IQR]: 0.57 [0.52-0.63] vs. 0.64 [0.58-0.81], p = 0.015). Delirious patients received higher doses of sedative and analgesic drugs, such as midazolam (median [IQR]: 3.82 [0.08-13.09] vs. 0 [0-0.56] mg/kg, p = 0.001), propofol (917 [496–1339] vs. 246 [0–562] mg/kg, p < 0.001), and fentanyl (343 [211–560] vs. 46 [0–177] mg/kg, p < 0.001). This resulted in a longer duration of coma (defined as a RASS of -5 or -4; median [IQR]: 12 [9–20] vs. 2 [0–7] days, p < 0.001) and longer ICU stays (median [IQR]: 20 [16–36] vs. 9 [5–12] days, p < 0.001).

Despite these differences, the use of antiviral and anti-inflammatory drugs was similar between the two groups, including steroids (46% vs. 35%, p = 0.541), tocilizumab (62% vs. 61%, p = 0.939), and hydroxychloroquine (48% vs. 52%, p = 0.792) (**Table 1**).

To account for the evolution of practices and standards of care during the COVID-19 pandemic, we compared patients admitted early (March and April 2020) to those admitted in a later phase (July 2020 to April 2021). The rate of delirium was non-significantly different between the two groups (56% vs. 73%, p = 0.361). Furthermore, most analyzed variables, including baseline characteristics, severity at admission, respiratory support, and doses of neuroleptic drugs, did not differ significantly (**Supp. Table 2**).

Reflecting the rapid evolution in understanding of COVID-19 and subsequent changes in the standard of care, we observed a significant shift from the use of hydroxychloroquine and tocilizumab to corticosteroids following results from the RECOVERY trial [18].

Patients’ sampling for multiplex bead assay and mass cytometry was performed after a median time of 2 days (2 [1–6] in delirious patients vs. 2 [1–8] in non-delirious ones, p=0.994) and before development of delirium in 35/39 patients (89%). The median time between sampling and first positive CAM-ICU was 16 days (IQR : 7-17).

### Immune signatures associated with COVID-19 and severity

We aimed to replicate the previously identified immune signature for COVID-19 patients within our cohort. To achieve this, we compared the levels of serum soluble markers in 62 ICU-admitted patients (ICU group) with 55 patients who did not require ICU hospitalization (non-ICU group) and 450 healthy donors (HD group) (**Fig. 1A**). COVID-19 patients, both ICU and non-ICU groups, exhibited a distinct immune signature compared to healthy donors (**Fig. 1B**). Consistent with previous studies, several cytokines (notably IL-6, IL-7, IL-1RA), chemokines (CXCL10, CXCL8, CXCL13), and growth factors (HGF, VEGF-A) were elevated in COVID-19 affected individuals (**Fig. 1C** and **Supp. Fig. 2**) [8, 17].

**Figure 1:**
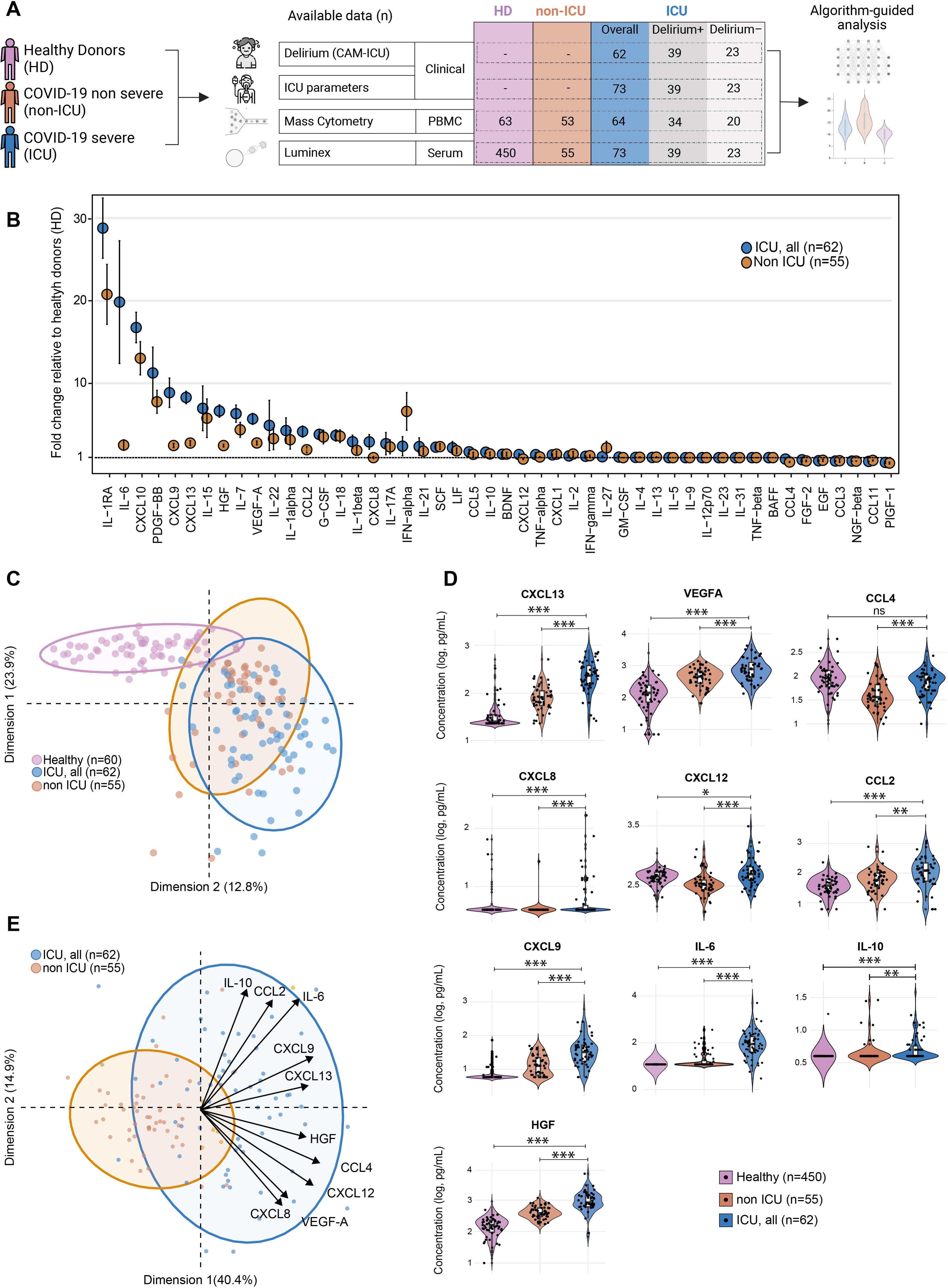
Pro-inflammatory signals correlate with COVID-19 severity. **(A)** Schematic overview of the study design. **(B)** Fold change in serum cytokine levels among COVID-19 patients requiring ICU care (ICU; blue dots, n=55), those not requiring ICU care (non-ICU; orange dots, n=62), and healthy donors (HD; n=450). Data are presented as median ± SEM.**(C)** Principal component analysis (PCA) comparing cytokine profiles across non-ICU patients (orange dots, n=62), ICU patients (blue dots, n=55), and healthy donors (violet dots, n=62). To improve visual clarity, the HD group was limited to a random subset of 62 samples from the total 450. **(D)** Box and violin plots comparing the concentrations of CXCL13, VEGF-A, CCL4, CXCL8, CXCL12, CCL2, CXCL9, IL-6, IL-10, and HGF in non-ICU (orange, n=62), ICU patients (blue, n=55), and healthy donors (violet, n=450), following log10 transformation. Additionally, individual data points (n=50 per group) are displayed as scatter plots, randomly selected from each group. Statistical significance was calculated using the Mann-Whitney U test on the full dataset: *p < 0.05, **p < 0.01, ***p < 0.001, ns: not significant (p > 0.05). **(E)** PCA illustrating the discriminatory power of the following cytokines: CXCL13, VEGF-A, CCL4, CXCL8, CXCL12, CCL2, CXCL9, IL-6, IL-10, and HGF, in differentiating severe (ICU; blue dots, n=55) from non-severe (non-ICU; orange dots, n=62) COVID-19 cases. The arrows indicate the relative contribution of each cytokine to the PCA model.

Additionally, we identified a set of serum soluble markers associated with disease severity and ICU admission among COVID-19 patients. These included CCL2, CCL4, CXCL9, IL-6, HGF, CXCL13, VEGF-A, IL-10, CXCL8, and CXCL12 (**Fig. 1D**), aligning with previous findings [8, 17]. This immune signature reliably differentiated between ICU and non-ICU groups (**Fig. 1E**).

We also analyzed the impact of immunomodulating treatments on severity-associated parameters among patients subsequently admitted to the ICU. Of these patients, 23 (37%) received corticosteroids before sampling, which was associated with a reduction in IL-6 levels and elevation of a set of peripheral chemokines/growth factors (CXCL8, CXCL12, CXCL13, VEGF-A) (**Supp. Fig. 3A**). In contrast, among the 36 patients (58%) who received tocilizumab, IL-6 levels were elevated, reflecting an increased soluble fraction due to IL-6 receptor blocking by this drug (**Supp. Fig. 3A)**. Given its immunomodulatory impact [19], we also analysed hydroxychloroquine. Similarly, IL-6 was elevated in patients receiving hydroxychloroquine before sampling (30 patients, 48%), though most of these patients also received tocilizumab (**Supp. Table. 1**).

### Serum Soluble Factors Associated with Delirium Partially Differ from Those Associated with Severity

To further characterize patients admitted to the ICU, we aimed to decipher how individual cytokines, chemokines, and growth factors correlate with key clinical endpoints associated with severity, including 30-day mortality, coma duration, need for mechanical ventilation, worst PaO_2_/FiO_2_ ratio, and severity (SAPSII) at admission. For instance, IL-1β and IL-15 levels were correlated with disease severity (SAPSII) at admission and with mortality after 30 days (**Fig. 2A**). Similarly, CXCL10, IL-22, IL-21, and IL-1α were negatively associated with the need for mechanical ventilation and coma duration (**Fig. 2A**). Additionally, the severity of hypoxia, measured by the worst PaO_2_/FiO_2_ ratio, was correlated with increased CXCL13 and decreased PDGF levels (**Fig. 2A**).

**Figure 2:**
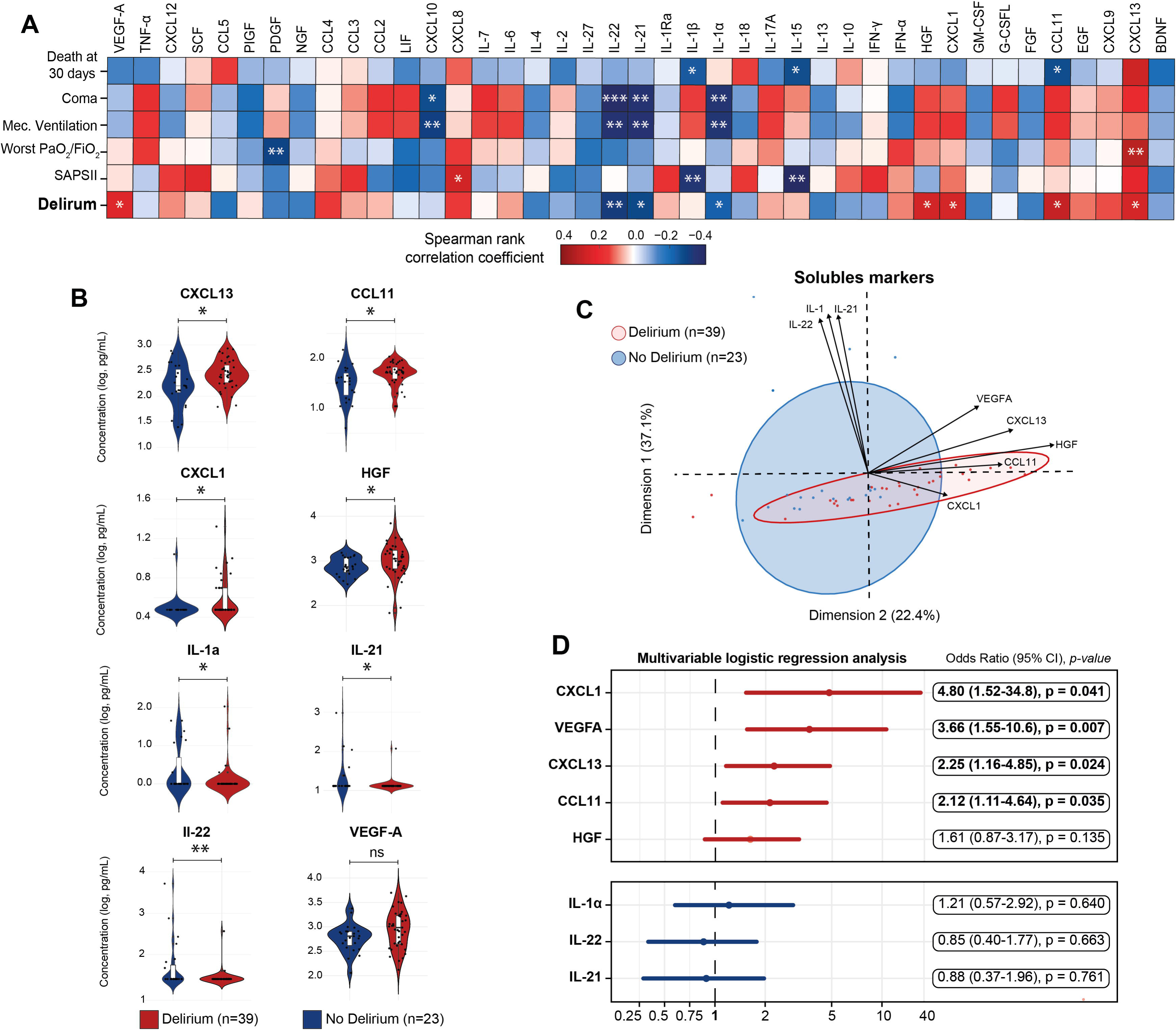
Serum-soluble markers independently correlate with the development of ICU-acquired delirium in COVID-19. **(A)** Correlation matrix between serum cytokines/chemokines/growth factors and clinical criteria, including 30-day mortality, coma, mechanical ventilation, worst PaOL/FiOL ratio, SAPS II score at admission, and delirium, in COVID-19 patients admitted to the ICU (n=62). Spearman’s rank correlation coefficients are represented by cell color, with significance levels denoted as follows: *p < 0.05; **p < 0.01; ***p < 0.001. **(B)** Box and violin plots comparing concentrations of soluble markers (after log10 transformation) in patients who developed delirium (red) vs. those who did not (blue). Markers showing significant correlations with delirium in the Spearman analysis include IL-1α, IL-21, IL-22, CXCL13, CCL11, CXCL1, HGF, and VEGF-A. Individual data points for each patient are displayed as scatter plots. Mann-Whitney U tests were used for statistical analysis, with significance represented as follows: *p < 0.05; **p < 0.01; ***p < 0.001; ns: not significant (p > 0.05).**(C)** Principal component analysis (PCA) showing the ability of selected cytokines (CCL11, CXCL1, CXCL13, VEGF-A, and HGF) to discriminate between patients with (red dots, n=39) and without (blue dots, n=23) delirium. IL-1α, IL-21, and IL-22 were excluded from the PCA due to minimal variation.**(D)** Multivariate logistic regression analysis of individual soluble markers (IL-1α, IL-21, IL-22, CXCL13, CCL11, CXCL1, HGF, and VEGF-A), adjusted for confounding factors including age, gender, worst PaOL/FiOL ratio, SAPS II score at admission, corticosteroid use, and doses of midazolam, propofol, and fentanyl. A forest plot depicts the odds ratios and 95% confidence intervals (CIs) for each marker. Markers positively correlated with delirium are shown in red (top panel), while those negatively correlated are shown in blue (bottom panel).

Overall, delirium was correlated with several cytokines (decreased IL-22, IL-21, IL-1α), chemokines (increased CXCL1, CCL11, CXCL13), and growth factors (increased HGF, and a tendency towards increased VEGF-A) (**Fig. 2A and B**). Interestingly, most of the severity markers described above (CCL2, CXCL12, CCL4, IL-10, CXCL8) were not associated with the development of delirium (**Fig. 2A**). Furthermore, most of these soluble markers were not affected by steroid administration prior to sampling, with the exception of CXCL13 and VEGF-A (**Supp. Fig. 4**). These markers were defining an immune signature differentiating patient s who would develop delirium (**Fig. 2C**).

We then analyzed the contribution of these markers while accounting for factors prone to favor delirium (age, gender, worst PaO_2_/FiO_2_ ratio, SAPSII score at admission, use of steroids, and analgo-sedation with fentanyl, midazolam, and propofol) using a multivariable logistic regression. This analysis demonstrated that only four serum soluble factors were independently associated with delirium: CXCL1, VEGF-A, CXCL13, and CCL11 (**Fig. 2D**).

### In depth immunophenotyping in delirious patients

To comprehensively characterize the inflammatory responses associated with delirium, we performed detailed immunophenotyping of circulating immune populations using mass cytometry (CyTOF). Utilizing an automated analysis with a pre-defined gating strategy, we evaluated 49 distinct cell populations.

Initially, we analyzed the correlation between specific immune populations and clinical endpoints. Certain cell types exhibited a statistically significant correlation predominantly with disease severity, including circulating follicular helper memory T cells, memory CD4+ T cells expressing PD1, naive CD8+ T cells, effector memory CD8+ T cells, conventional dendritic cells, transitional B cells, class-switched memory B cells (IgG2), and CD10+ transitional B cells (T1 B Cells). Other cell types showed significant correlations primarily with delirium, such as memory CD4+ T cells expressing CXCR3, activated T cells, IgM+ unswitched memory B cells, and exhausted B cells (CD21^low^ CD38l^ow^) (**Fig. 3A**). However, using univariate analysis, only an increase in exhausted B cells and a decrease in activated T cells were significantly associated with the development of delirium (**Fig. 3B**). These cellular population were poorly able to discriminate between groups (**Fig. 3C**).

**Figure 3:**
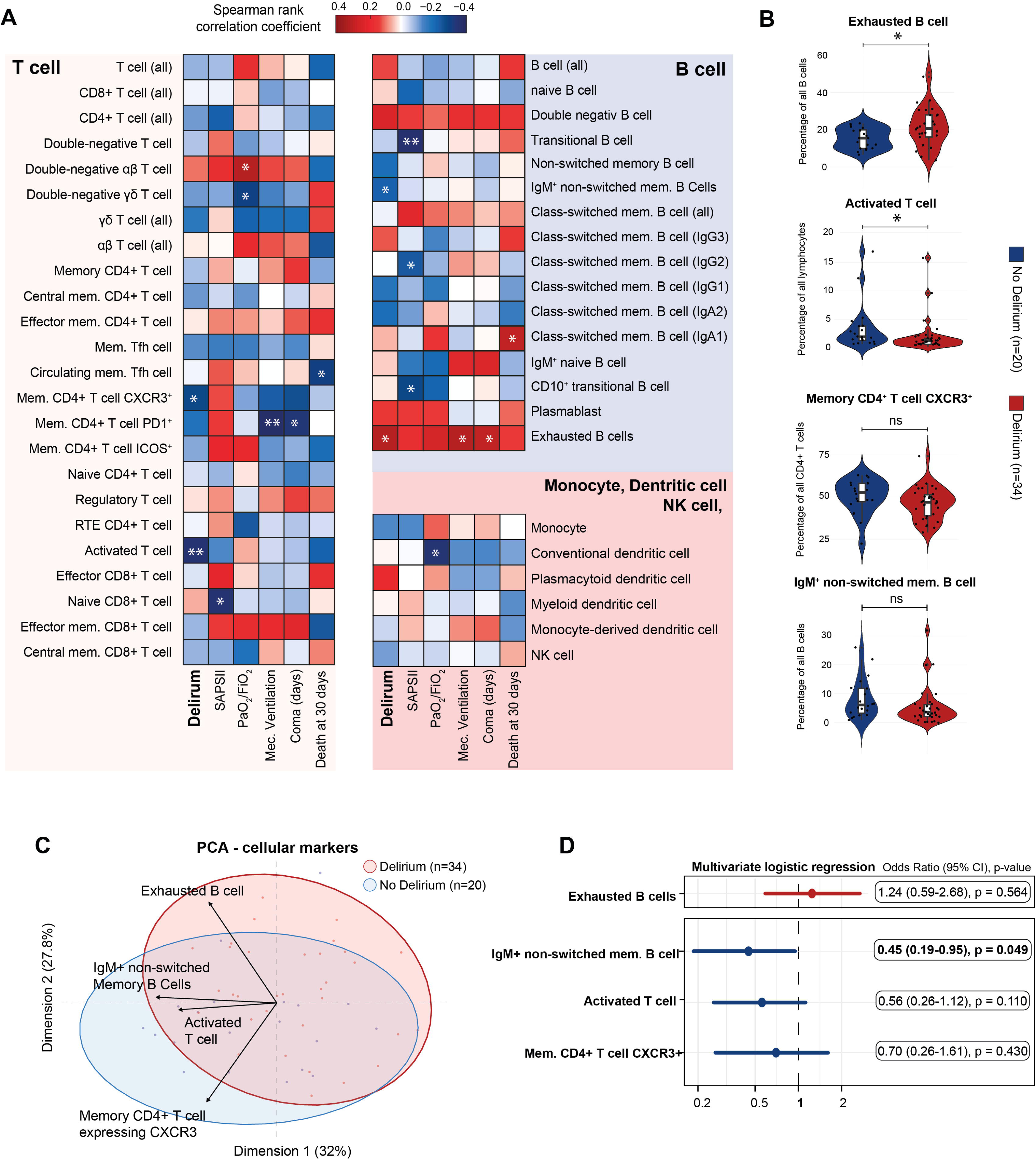
Immunophenotyping reveals a specific B cell population associated with delirium. **(A)** Correlation matrix between immune populations and clinical criteria, including 30-day mortality, coma, mechanical ventilation, worst PaOL/FiOL ratio, SAPS II score at admission, and delirium, in COVID-19 patients admitted to the ICU (n=62). Spearman’s rank correlation coefficients are represented by cell color, with significance levels denoted as follows: *p < 0.05; **p < 0.01; ***p < 0.001. **(B)** Box and violin plots comparing concentrations of cell population (after log10 transformation) in patients who developed delirium (red) vs. those who did not (blue). Markers showing significant correlations with delirium in the Spearman analysis include exhausted B cell, IgM^+^ non-switched memory B-cell, activated T cell, and memory CD4^+^ T cell expressing CXCR3. Individual data points for each patient are displayed as scatter plots. Mann-Whitney U tests were used for statistical analysis, with significance represented as follows: *p < 0.05; **p < 0.01; ***p < 0.001; ns: not significant (p > 0.05).**(C)** Principal component analysis (PCA) showing the poor ability of selected cytokines (exhausted B cell, IgM^+^ non-switched memory B-cell, activated T cell, and memory CD4^+^ T cell expressing CXCR3) to discriminate between patients with (red dots, n=39) and without (blue dots, n=23) delirium. **(D)** Multivariate logistic regression analysis of individual cellular population (exhausted B cell, IgM^+^ non-switched memory B-cell, activated T cell, and memory CD4^+^ T cell expressing CXCR3), adjusted for confounding factors including age, gender, worst PaOL/FiOL ratio, SAPS II score at admission, corticosteroid use, and doses of midazolam, propofol, and fentanyl. A forest plot depicts the odds ratios and 95% confidence intervals (CIs) for each marker. Markers positively correlated with delirium are shown in red (top panel), while those negatively correlated are shown in blue (bottom panel).

Subsequently, we investigated whether the cell types previously correlated with delirium (memory CD4+ T cells expressing CXCR3, activated T cells, IgM+ unswitched memory B cells, and exhausted B cells) remained independently associated with delirium after adjusting for known confounding factors (age, gender, worst PaO_2_/FiO_2_ ratio, SAPSII score at admission, use of corticosteroids, doses of midazolam, doses of propofol, and doses of fentanyl). We employed multivariable logistic regression models to test each cell type independently. The association remained statistically significant only for IgM+ non-switched memory B cells (**Fig. 3D**).

### Prognostic Immune Signatures for Delirium

To explore whether immune parameters could predict delirium development, we developed a logistic regression model using L2 regularization (ridge regression) due to the high number of parameters (p = 83) relative to our sample size (n = 54). The dataset was split into a training set (80%) and a test set (20%) (**Fig. 4A**). We tested five models: one with all serum soluble factors, one with blood phenotyping data, one combining both, and one including the 12 parameters previously associated with delirium (IL-1α, IL-22, IL-21, CCL11, CXCL1, CXCL13, HGF, VEGF-A, CXCR3+ CD4+ T cells, IgM+ unswitched memory B cells, exhausted B cells, HLA-DR+ activated T cells). The best AUC-ROC was achieved with the latter (**Fig. 4B**).

**Figure 4.**
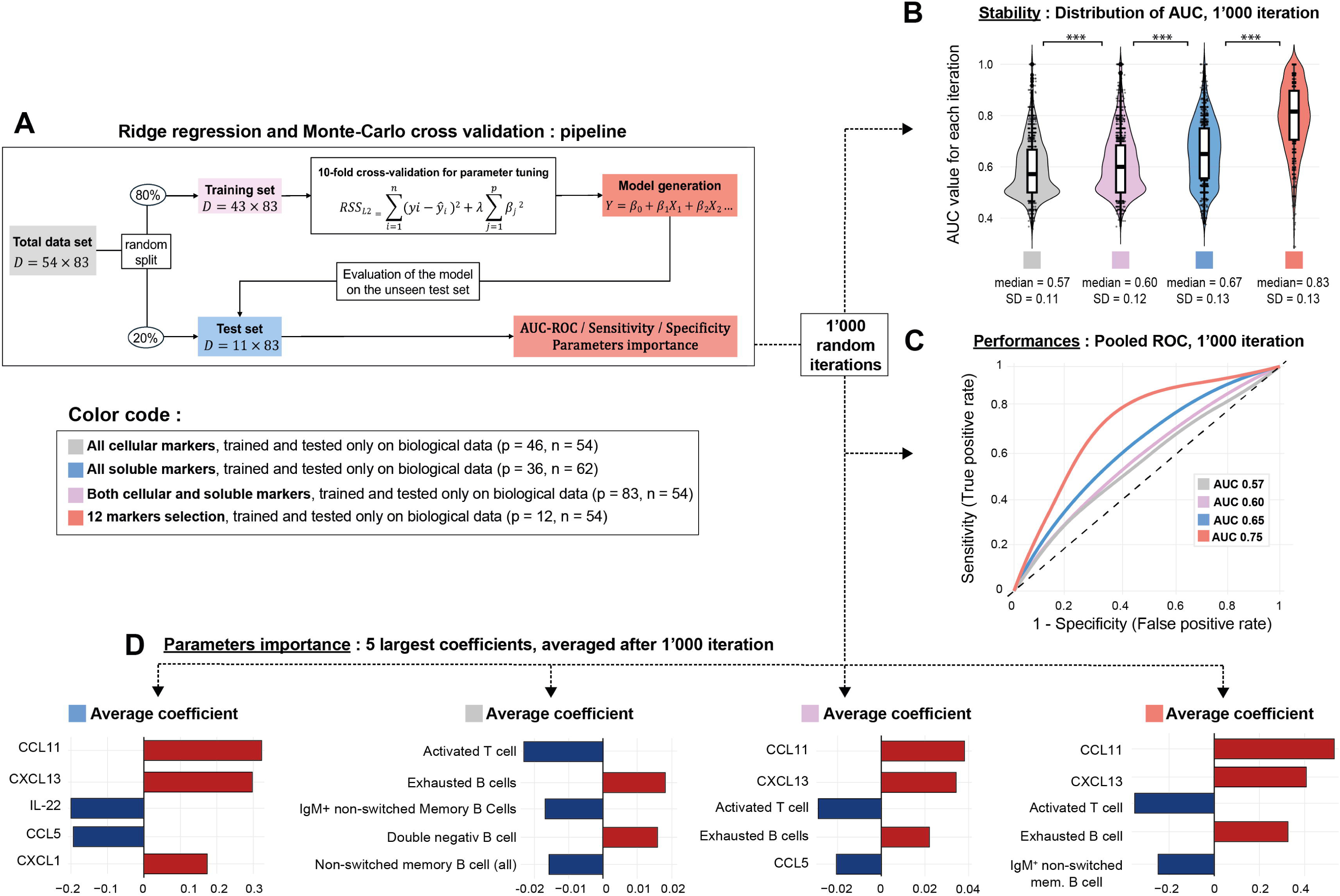
Soluble markers and/or immunophenotyping for prediction of ICU-acquired delirium. **(A)** Schematic of the analysis pipeline using the combined dataset (p = 83, n = 54) as an example. The same approach was applied to datasets of different dimensions. **(B)** Box and violin plots showing accuracy and variability in AUC across 1’000 iterations for each model: all cellular markers (p = 46, n = 54; gray; median = 0.57, SD = 0.11), all soluble markers (p = 36, n = 62; blue), both cellular and soluble markers (p = 84, n = 54; pink), and the 12 markers previously linked to delirium (p = 84, n = 54; red). Individual data points from each iteration are overlaid as scatter plots. Mann-Whitney U tests were used for statistical comparisons (***p < 0.001). **(C)** ROC curves depicting the overall poor performance of models across the four datasets after pooling AUC from 1,000 iterations. All cellular markers (gray), all soluble markers (blue), both cellular and soluble markers (pink), and the 12 markers previously linked to delirium (red). **(D)** Histogram of averaged coefficients from 1’000 iterations, highlighting the top five features for each dataset. Blue bars indicate positive associations with delirium, while red bars show negative associations. From left to right: top five features for models based on soluble markers, cellular markers, combined markers, and the 12 markers previously associated with delirium. Abbreviation: *D* = Dimension of the matrix; RSS_L2_ = residual sum of squares; Y_i_ = observed value; Ȳ = predicted value; λ= penalty parameter; *β_j_* = coefficient; AUC-ROC = area under the curve of the receiver operating curve.

We report here the area under curve (AUC) of the pooled receiving operator curves (ROC) for 1 000 random iterations testing the model. The first model, using only soluble markers (36 parameters, n = 62), claimed a mean AUC-ROC of 0.57 (**Fig. 4C and D**), with CCL11, CXCL13, and IL-22 being the most important parameters. The second model, using only immunophenotyping data (46 parameters, n = 54), showed a mean AUC-ROC of 0.60, with activated T cells and exhausted B cells as key contributors (**Fig. 4C and D**). The third model, combining all markers (83 parameters, n = 54), achieved a mean AUC-ROC of 0.65 (**Fig. 4C and D**). Finally, the model with the 12 key markers had a mean AUC-ROC of 0.75 (**Fig. 4C and D**), indicating that a small set of biomarkers can provide a better predictive power.

Due to the limited sample size that prevented us from training our model correctly and achieving stable AUC-ROC, we generated synthetic data using beta-variational autoencoders. We carefully tuned the parameters to minimize pollution from the original data, ensuring the synthetic samples were safe for downstream training tasks. This increased our total sample size to 1’000 (**Fig. 5A**). We selected once again the 12 parameters previously associated with delirium (p = 12, n = 1’000) and reapplied the same pipeline. This time, the AUC-ROC generated at each iteration was much more stable (**Fig. 5B**), and we achieved a pooled AUC-ROC of 0.89 (**Fig. 5C**). The best model achieved an AUC of 0.95, a sensitivity of 93%, a specificity of 86%, a positive predictive value (PPV) of 89%, and a negative predictive value (NPV) of 86% (**Fig. 5D**).

**Figure 5.**
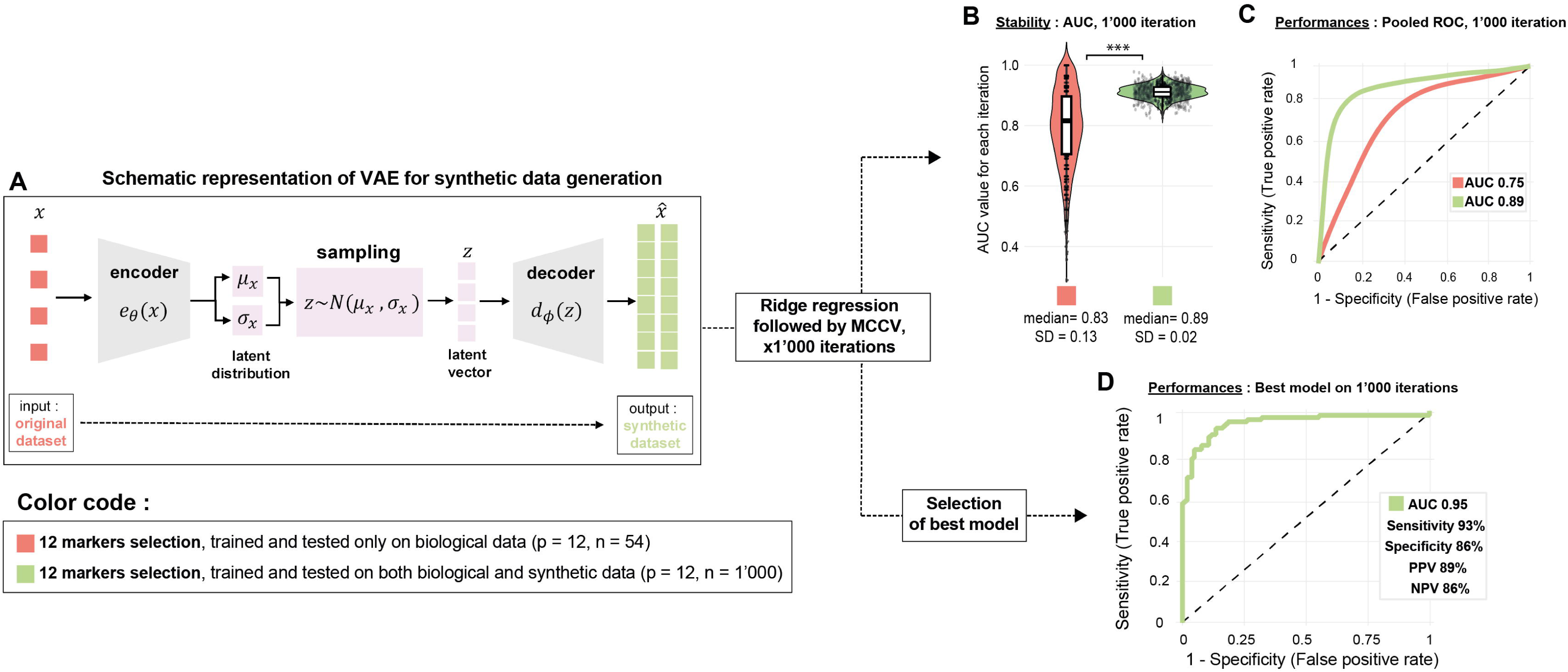
The utilization of synthetic data enhances model stability and performance to predict delirium. **(A)** Schematic representation of the variational autoencoders employed for generating synthetic data. (B) Box and violin plots illustrate the accuracy and variability in the area under the curve (AUC) across 1,000 iterations for models trained on both biological and synthetic data (green, n = 1,000) compared to models trained exclusively on biological data (red, n = 54). Individual data points for each iteration are overlaid as scatter plots. Mann-Whitney U tests were performed for statistical comparisons (***p < 0.001). (C) Receiver operating characteristic (ROC) curves of models incorporating both biological and synthetic data (green) versus those using only biological data (red;), derived from pooling AUC values across 1,000 iterations. (D) ROC curve obtained with the optimal ridge regression model achieved an AUC of 0.95, with performance metrics sensitivity, specificity, positive predictive value (PPV), and negative predictive value (NPV).

## Discussion

In this study, we aimed to dissect the inflammatory environment associated to delirium in ICU patients with ARDS due to severe SARS-CoV-2 infection. We identified several soluble factors and immune cell types linked to delirium, including IL-1α, IL-22, IL-21, CCL11, CXCL1, CXCL13, and VEGF-A, as well as exhausted B cells and activated T cells. Notably, CCL11, CXCL1, CXCL13, and VEGF-A remained significantly associated with delirium after adjusting for confounding factors. Using these markers, we developed a predictive model for delirium.

While CXCL1 has been previously associated with delirium [20, 21], associations with CCL11, CXCL13, and VEGF-A are novel findings, likely due to limited prior investigations of these markers in delirium [12]. However, their roles in BBB dysfunction and cognitive impairment have previously been suggested, including in COVID-19 patients [9, 11]. Then, VEGF-A, CXCL13/CXCR5 axis and CXCL1/CXCR2 axes have been shown to contribute to BBB dysfunction and promote neuroinflammation [22–25]. Of note, VEGF-A has been linked to both cognitive impairment (HIV associated neurological disorders) [26] and neuroprotection (Alzheimer disease) [27], suggesting a complex role in neuroinflammation. Similarly, CXCR5, the receptor for CXCL13, is linked to hippocampal neuroinflammation and cognitive impairment in animal models of sepsis [28] and surgery [29].

Furthermore, CCL11, associated with aging and cognitive decline, has been implicated in reduced neurogenesis and synaptic density, notably in Alzheimer’s disease models [30, 31]. COVID-19 survivors with cognitive impairment show higher levels of CCL11, supporting its role in COVID-related cognitive dysfunction [32].Last, we have shown a reduction, among other cytokines, of IL-22. This mediator have been shown also to be involved at the BBB interface and to promote astrocytes survival [33].

Regarding immune cells, exhausted B cells (CD21^low^) are hallmark of chronic activation within the adaptive immune system and has been observed in various context, including infections (i.e. HIV and HCV associated), immunodeficiency (i.e. common variable immunodeficiency) and autoimmune disorders (i.e. systemic lupus erythematosus and rheumatoid arthritis). Notably, an elevated proportion of CD21^low^ B cells has been reported in cases of acute immune dysregulation, including septic shock [34, 35] and severe COVID-19 [36]. In our cohort of patients with delirium, we also observed a reduction of activated HLA-DR+ T lymphocytes expressing, a finding that could potentially also reflect underlying immune exhaustion. Indeed, reduced HLA-DR expression has previously been documented in sepsis, where it is associated with immunosuppression and worse clinical outcomes, including increased mortality [37].

Taken together, these findings indicate that delirium might arise from the conjunction of both local blood-brain barrier disruptions and systemic immune dysregulation, though further investigation is needed to clarify the pathophysiological mechanisms at the root of delirium.

In addition to their role in the pathophysiology of infection-related delirium, we demonstrated that a targeted set of cellular and soluble markers may predict delirium development with high accuracy (AUC 0.95). Our biomarker-based model potentially outperforms current clinical prediction models, such as PRE-DELIRIC and E-PRE-DELIRIC, which show AUCs ranging from 0.68 to 0.87 and 0.68 to 0.72, respectively [38–40]. Moreover, using biomarkers provides a dynamic opportunity to monitor patient evolution in the ICU setting. Identifying patients at high risk for delirium could enable targeted interventions, such as reducing sedative use and increasing non-pharmacological measures, to prevent its onset [16, 41].

Our study has several limitations. First, its retrospective design, small sample size, and the absence of long-term follow-up beyond the ICU setting are important constraints. Additionally, limiting the analysis to COVID-19 patients restricts the generalizability to all cases of infection- or sepsis-associated delirium, as well as delirium related to other etiologies. However, it has been shown that immune signatures in COVID-19 share similarities with those in other infections, suggesting some overlap [8].

These results need to be validated in a prospective study, incorporating a broader range of etiologies underlying delirium. In such a setting, it would be crucial to compare the performance of our model against existing models, such as PRE-DELIRIC, and assess whether combining them could enhance predictive accuracy. Additionally, exploring the potential of integrating other biomarkers, such as those reflecting neuronal damage (NfL) or astroglial activation (GFAP) [42], along with electrophysiological parameters [43], could improve prediction and capture the dynamic evolution of delirium. It would also be crucial to identify patients at risk of developing cognitive disorders following delirium [2, 3].

## Declarations

### Ethics approval and consent to participate

This study was approved by the local Ethics Committee (CER-VD) under the CORO-NEURO study (authorization n°. 2020-01123). Informed consent was obtained from all living participants; however, it was waived for those who had passed away.

### Authors Contributions

TB and RBV contributed to the study design, data analysis, interpretation, and drafting of the manuscript. PPA and AP participated in data analysis and critical revision of the manuscript. MP, NBH, and CF were responsible for data acquisition and manuscript revision. NBH, AOR, JDC and RDP critically revised the manuscript.

## Acknowledgements.

We would like to thank Nadine Do Rosario and Nathalie Felix for their technical help.

