## Supplementary Appendix for "Unravelling the Complex Inflammatory Landscape of COVID-19 infection: A Pathway to Biomarkers Identification in Infection-Associated Delirium in the ICU"

*Title:*

Dr. Raphaël Bernard-Valnet

Centre hospitalier universitaire Vaudois (CHUV), Rue du Bugnon 46, 1011 Lausanne

**Supplementary Material and Methods**

**Assessment of serum immune profile**

Serum concentrations of various cytokines, soluble cytokine receptors, chemokines, and growth factors. Here is the list of cytokines and growth factors included in this study: interleukin-1 alpha (IL-1α), interleukin-1 receptor antagonist (IL-1RA), interleukin-1 beta (IL-1β), interleukin-2 (IL-2), interleukin-4 (IL-4), interleukin-5 (IL-5), interleukin-6 (IL-6), interleukin-7 (IL-7), interleukin-9 (IL-9), interleukin-10 (IL-10), interleukin-12p70 (IL-12p70), interleukin-13 (IL-13), interleukin-15 (IL-15), interleukin-17A (IL-17A), interleukin-18 (IL-18), interleukin-21 (IL-21), interleukin-22 (IL-22), interleukin-23 (IL-23), interleukin-27 (IL-27), interleukin-31 (IL-31), interferon-alpha (IFN-α), interferon-gamma (IFN-γ), tumor necrosis factor alpha (TNFα), chemokines including C-C motif chemokine ligand 2 (CCL2), C-C motif chemokine ligand 3 (CCL3), C-C motif chemokine ligand 4 (CCL4), C-C motif chemokine ligand 5 (CCL5), C-C motif chemokine ligand 11 (CCL11), C-X-C motif chemokine ligand 1 (CXCL1), C-X-C motif chemokine ligand 8 (CXCL8), C-X-C motif chemokine ligand 9 (CXCL9), C-X-C motif chemokine ligand 10 (CXCL10), C-X-C motif chemokine ligand 12 (CXCL12), C-X-C motif chemokine ligand 13 (CXCL13), tumor necrosis factor-beta (TNF-β), nerve growth factor-beta (NGF-β), brain-derived neurotrophic factor (BDNF), epidermal growth factor (EGF), fibroblast growth factor-2 (FGF-2), hepatocyte growth factor (HGF), leukemia inhibitory factor (LIF), platelet-derived growth factor-BB (PDGF-BB), placental growth factor-1 (PlGF-1), stem cell factor (SCF), vascular endothelial growth factor-A (VEGF-A), vascular endothelial growth factor-D (VEGF-D), B-cell activating factor (BAFF), granulocyte-macrophage colony-stimulating factor (GM-CSF), and granulocyte colony-stimulating factor (G-CSF) (n = 49). There were determined using a multiplex bead assay (Luminex assay - ProcartaPlex Mix&Match Human plex, Thermofischer) as previously described ^14^.

**Immunophenotyping by mass cytometry**

The blood concentration and relative proportions of different CD4 and CD8 T-cell populations, B-cell populations, gamma-delta T cells, NK cells, monocytes, and dendritic cell populations were assessed by mass cytometry using a panel of 45 surface markers as previously described ^14^. The following antibodies were used for mass cytometry experiments. Panel 1: 111Cd-conjugated anti-CD141 (1A4), 113In-conjugated anti-CD8 (RPA-T8), 115In- conjugated anti-CD4 (RPA-T4), 116Cd-conjugated anti-IgA2 (A9604D2), 141Pr-conjugated anti-CD45 (HI30), 142Nd-conjugated anti-CD19 (HIB19), 143Nd-conjugated anti-ICOS (C398.4A), 144Nd-conjugated anti-IgG3 (HP6047), 145Nd-conjugated anti-CD31/PECAM-1 (WM59), 146Nd-conjugated anti-IgD (IA6-2), 147Sm-conjugated anti-CD7 (CD7-6B7), 148Nd-conjugated anti-IgA1 (B3506B4), 149Sm-conjugated anti-CD127 (A019D5), 150Nd- conjugated anti-IgG1 (G17-1), 151Eu-conjugated anti-CD123 (6H6), 152Sm-conjugated anti- CD21 (BL13), 153Eu-conjugated anti-CD62L (DREG-56), 154Sm-conjugated anti-CD3 (UCHT1), 155Gd-conjugated anti-CD27 (L128), 156Gd-conjugated anti-TCR γ (gamma)/δ (delta) (B1), 158Gd-conjugated anti-CD10 (HI10a), 159Tb-conjugated anti-CD197/CCR7 (G043H7), 160Gd-conjugated anti-CD14 (M5E2), 161Dy-conjugated anti-CD1c (L161), 162Dy-conjugated anti-CD11c (Bu15), 163Dy-conjugated anti-CD183/CXCR3 (G025H7), 164Dy-conjugated anti-CD185/CXCR5 (51505), 165Ho-conjugated anti-CD45RO (UCHL1), 166Er-conjugated anti-CD24 (ML5), 167Er -conjugated anti-CD38 (HIT2), 168Er- conjugated anti-CD66b (G10F5), 169Tm-conjugated anti-CD25 (2A3), 170Er-conjugated anti-CD45RA (HI100), 171Yb-conjugated anti-CD20 (2H7), 172Yb-conjugated anti-IgM (MHM-88), 173Yb-conjugated anti-TCR α(alpha)/ β(beta) (T1089.A-31), 174Yb-conjugated anti-HLA-DR (L243), 175Lu-conjugated anti-CD279/PD-1 (EH12.2H7), 176Yb-conjugated anti-CD56 (HCD56), 198Pt-conjugated anti-IgG2 (HP6002), 209Bi-conjugated anti-CD16 (3G8), 112Cd-conjugated anti-CD69 (FN50), 106Cd-conjugated anti-CCR6 (11A9), 194Pt- conjugated anti-CCR4 (L291H4) and 191Ir was used to label DNA. Antibodies against TCR α(alpha)/ β(beta), IgG1, CD66b, CCR6 and CD141 were purchased from BD. Antibodies against TCR γ (gamma)/δ (delta), CD278/ICOS, IgG2, IgG3, CD1c, CD4, CD8, CD69 and CCR4 were purchased from Biolegend. Antibodies against IgA1 and IgA2 were purchased from SouthernBiotech. All were conjugated with Maxpar® X8 Antibody Labeling Kit except IgA2, CD141, CD69 and CCR6 who were labelled using Maxpar MCP9 Antibody Labeling Kit. All other antibodies were purchased from Fluidigm/DVS. DNA positive cells were assessed using Cell-ID Intercalator-Ir (#201192B) from fluidigm. Panel 2: 113In-conjugated anti-CD8 (RPA-T8), 115In-conjugated anti-CD4 (RPA-T4), 149Sm-conjugated anti-CCR4 (L291H4), 176Yb-conjugated anti-CD127 (A019D5), 141Pr-conjugated anti-CCR6 (G034E3), 154Sm-conjugated anti-CXCR3 (G025H7), 168Er-conjugated anti-CCR9 (L053E8), 159Tb-conjugated anti-CCR7 (G043H7), 167Er-conjugated anti-CXCR5 (J252D4), 144Nd-conjugated anti-CCR5 (NP-6G4), 106Cd-conjugated anti-CD45 (HI30), 111Cd-conjugated, anti-CD3 (UCHT1), 142Nd-conjugated anti-CD44 (IM7), 158Gd-conjugated anti-CD25 (M- A251), 141Pr-conjugated anti-CCR6 (G034E3), 163Dy-conjugated anti-CD38 (HIT2), 153Eu- conjugated anti-TIGIT (MBSA43), 147Sm-conjugated anti-2B4 (C1.7), 151Eu-conjugated anti-PD1 (EH12.2H7), 155Gd-conjugated anti-CD27 (L128), 162Dy-conjugated anti-CD69 (FN50), 164Dy-conjugated anti-CD45RO (UCHL1), 209Bi-conjugated anti-CD16 (3G8), 145Nd-conjugated anti-CD31 (WM59), 161Dy-conjugated anti-CD95 (DX2), 194Pt- conjugated anti-CD57 (NK-1), 166Er-conjugated anti-NKG2D (ON72), 170Er-conjugated anti-CD45RA (HI100), 174Yb-conjugated anti-HLADR (L243), 148Nd-conjugated anti- PDL1 (29E.2A3), 171Yb-conjugated anti-CD151 (50/6), 152Sm-conjugated anti-CD40L (TRAP1), 143Nd-conjugated anti-ICOS (C398.4A), 172Yb-conjugated anti-LAG3 (874501), 150Nd-conjugated anti-OX40 (ACT35), 160Gd-conjugated anti-Tbet (4B10), 165Ho- conjugated anti-Ki67 (Ki67), 169Tm-conjugated anti-Bcl2 (100), 175Lu-conjugated anti- RorγT (AFKJS-9), 146Nd-conjugated anti-Gata3 (TWAJ), 156Gd-conjugated anti-FoxP3 (PCH101) and 191Ir was used to label DNA. Antibodies against CD45, CD8, CD4, CD44, ICOS, 2B4, PD-1, CXCR3, CD25, CCR7, CD38, Ki67, CXCR5, Bcl2 were purchased from Biolegend. Antibodies against CD3, CD40L (CD154), CD57 and CD151 were purchased from BD. Antibodies against Gata3, FoxP3, CD95(FAS) and RorγT were purchased from e- Biosciences. Antibody against LAG3 was purchased from R&D Systems. All were conjugated with Maxpar® X8 Antibody Labeling Kit except CD45 and CD3 who were labelled using Maxpar MCP9 Antibody Labeling Kit. All other antibodies were purchased from Fluidigm/DVS. DNA positive cells were assessed using Cell-ID Intercalator-Ir (#201192B) from fluidigm. Panel 3: 154Sm -conjugated anti-CD3 (UCHT1), 106Cd -conjugated anti-CD45 (HI30), 113In -conjugated anti-CD8 (RPA-T8), 115In -conjugated anti-CD4 (RPA-T4), 142Nd -conjugated anti-CD19 (HIB19), 143Nd -conjugated anti-CD1c (L161) , 144Nd -conjugated anti-CD69 (FN50), 145Nd -conjugated anti-CD31 (WM59), 146Nd -conjugated anti-CD86 (GL-1) , 147Sm -conjugated anti-CD7 (CD7-6B7), 148Nd -conjugated anti-CD39 (A1) , 149Sm -conjugated anti-CD56 (HCD56), 150Nd -conjugated anti-pSTAT5 (47), 151Eu- conjugated anti-CD123 (6H6), 152Sm -conjugated anti-CD21 (BL13), 153Eu -conjugated anti- pSTAT1 [Y701] (58D6), 155Gd -conjugated anti-CD27 (L128), 156Gd -conjugated anti-p38 [T180/Y182] (D3F9), 158Gd -conjugated anti-pSTAT3 (4/P-Stat3), 159Tb -conjugated anti- pMAPKAPK2 (27B7), 160Gd -conjugated anti-CD14 (M5E2), 162Dy -conjugated anti- CD11c (Bu15), 163Dy -conjugated anti-CD62L (DREG-56) , 164Dy -conjugated anti-CD161 (HP3G10), 165Ho -conjugated anti-pNFkb (K10895.12.50) , 166Er -conjugated anti-CD20 (2H7), 167Er -conjugated anti-CD38 (HIT2), 168Er -conjugated anti-Ki67 (Ki67) , 169Tm -conjugated anti-CD45RA (HI100) , 171Yb -conjugated anti-pERK1/2 [T202/Y204] (D13.14.4E), 172Yb -conjugated anti-CD15 [SSEA-1] (W6D3), 173Yb -conjugated anti- CD141 (1A4), 174Yb -conjugated anti-HLA-DR (L243), 175Lu -conjugated anti-pS6 (N7548), 176Yb -conjugated anti-pCREB (87G3), 194Pt -conjugated anti-CD57 (NK-1), 209Bi -conjugated anti-CD16 (3G8) and 191Ir was used to label DNA. Antibodies against Ki67, CD45, CD8a, CD4, CD1c, CD69, CD86, CD39, CD56, CD62L and CD45RA were purchased from Biolegend. Antibodies against NF-kB p65, CD20, CD141 and CD57 were purchased from BD. All were conjugated with Maxpar® X8 Antibody Labeling Kit except CD45 which was labelled using Maxpar MCP9 Antibody Labeling Kit. All other antibodies were purchased from Fluidigm/DVS. DNA positive cells were assessed using Cell-ID Intercalator-Ir (#201192B) from fluidigm.

The gating strategy also have been extensively described in a previous publication [17]..

**Statistical Analysis**

To account for possible confounding factors while analyzing biomarkers associated with delirium, we also conducted multivariate binomial logistic regressions considering age, sex, SAPS II, worst PaO2/FiO2 ratio, use of corticosteroids, and the doses of midazolam, propofol, and fentanyl received. Additionally, we employed principal component analysis (PCA) as a linear dimensionality reduction tool to visualize our dataset.

Plot generation and statistical analyses were performed using R statistical software (v4.4.0; R Core Team 2023) and the following additional packages: readxl (for importing Excel files), ggplot2 (for plot generation), glmnet (for multivariate regression model and ridge regression model generation), pROC (for ROC curve generation and analysis), and rstatix (for performing statistical tests).

**Prediction models.**

We utilized a ridge regression model to optimize the variable selection process and create an effective model for predicting delirium based on immunological biomarkers. First, we remove all parameters without variation between the 2 groups. Ridge regression adds a penalty proportional to the square of the magnitude of the coefficients ($\beta_{j}$) and the penalty parameter $\lambda$. The model is fitted by trying to minimize the residual sum of squares (${RSS}_{L2}$), which can be understood as the addition of the inaccuracy of the model (square of the difference between the predicted and the observed output) and a penalty that increases with the model complexity and the size of the coefficients. This shrinks the coefficient values towards zero, helping to prevent overfitting by creating a simpler and sparser model. Doing so optimizes the bias-variance tradeoff by increasing bias but further reducing variance, resulting in a model that performs better on new, unseen data.

To build the model, we split the dataset randomly into a training set (80% of the dataset) and a test set (remaining 20%). We used 10-fold cross-validation on the training set to tune the ridge regression parameters, including the penatly parameter $\lambda$ and the coefficients $\beta_{j}$ associated with each variable. After fitting the model, we evaluated its performance on the unseen test set by calculating the area under the receiver operating characteristic curve (AUC-ROC), sensitivity, and specificity. Additionally, we identified the most important parameters, indicated by the largest absolute values of the coefficients $\beta_{j}$.

Due to the risk of inaccuracy in evaluating the model's performance on unseen data caused by the random split of a small dataset into a training and a test sets, we incorporated an additional validation step. For each tested dataset, we trained and tested 1’000 slightly different models and extracted the AUC-ROC after each iteration to assess performance reproducibility across different splits, a technique known as Monte Carlo Cross-Validation (or repeated random subsampling validation). Indeed, with an 80-20 split of 54 samples, there are more than 12 billion possible combinations, some of which are expected to overestimate and others to underestimate the real-life performance of the trained model (that is, on unseen data outside of the dataset). Results were represented as a receiver operating characteristic (ROC) curve with the associated area under the curve (AUC), sensitivity, and specificity.

We employed a BetaVAE (β-Variational Autoencoder) to generate synthetic data. BetaVAE incorporates a β term that controls the balance between reconstruction loss and KL divergence, allowing for a trade-off between data fidelity and latent space exploration. The model encodes the data into a latent space, reparametrizes it using a Gaussian distribution, and then decodes it back into the original space. Two separate model instances were trained for positive and negative samples, using the Adam optimizer to minimize both reconstruction error and KL divergence. This approach enabled the generation of realistic synthetic samples while maintaining a careful balance between data accuracy and latent space variability. To ensure the synthetic data's integrity and safety for downstream training tasks, we carefully tuned the model parameters to minimize contamination from the original dataset.

**Large Language Model.** ChatGPT 4.0 (OpenAI) has been used for English proofreading and for code generation/correction for R.

### **Supplementary Tables and Figures**

**Supplementary Table 1 -** Correspondance between cell populations name and surface markers assessed.

| Cell Surface Marker | Cell name |
| --- | --- |
| *Monocyte, Dendritic cell, NK cell* |  |
| CD14^+^ | Monocyte |
| HLA-DR^+^ CD64^+^ | Monocyte-derived dendritic cell (Mo-DC) |
| CD14^–^ HLA-DR^+^ CD11c^+^ | Myeloid dendritic cell (mDC) |
| CD14^–^ HLA-DR^+^ CD11c^+^ CD1c^+^ | Conventional dendritic cell type 2 (cDC2) |
| CD14^–^ HLA-DR^+^ CD141^+^ | Conventional dendritic cell type 1 (cDC1) |
| CD14^–^ HLA-DR^+^ CD123^+^ | Plasmacytoid DC (pDC) |
| CD56^+^ CD3^–^ | Lymphocytes NK |
| *T cell* |  |
| CD3^+^ | T cell (all) |
| CD3^+^ HLD-DR^+^ | Activated T cell |
| CD3^+^ CD4^–^ CD8^–^ | Double-negative T cell (all) |
| CD3^+^ γδ-TCR^+^ | Gamma delta T cell (all) |
| CD3^+^ CD4^–^ CD8^–^ γδ-TCR | Double-negative gamma delta T cell |
| CD3^+^ αβ-TCR | Alpha beta T cell (all) |
| CD3^+^ CD4^–^ CD8^–^ αβ-TCR | Double-negative alpha beta T cell |
| CD3^+^ CD4^+^ | CD4^+^ T cell (all) |
| CD3^+^ CD4^+^ CCR7^+^ | Naive CD4^+^ T cell |
| CD3^+^ CD4^+^ CD45RA^+^ CD31^+^ | Recent thymic emigrant CD4^+^ T cell |
| CD3^+^ CD4^+^ CD45RA^–^ | Memory CD4^+^ T cell |
| CD3^+^ CD4^+^ CD45RA^–^ CCR7^–^ | Effector memory CD4+ T cell |
| CD3^+^ CD4^+^ CD45RA^–^ CCR7^+^ | Central memory CD4+ T cell |
| CD3^+^ CD4^+^ CD45RA^–^ CXCR3^+^ | Memory CD4+ T cell expressing CXCR3 |
| CD3^+^ CD4^+^ CD45RA^–^ PD1^+^ | Memory CD4+ T cell expressing PD1 |
| CD3^+^ CD4^+^ CD45RA^–^ ICOS^+^ | Memory CD4+ T cell expressing ICOS |
| CD3^+^ CD4^+^ CD45RA^–^ CXCR5^+^ | Follicular helper memory cell |
| CD3^+^ CD4^+^ CD45RA^–^ CXCR5^+^ PD1^+^ ICOS^+^ CXCR3^–^ | Circulating memory follicular helper T cell |
| CD3^+^ CD4^+^ CD25^+^ CD127^–^ | Regulatory T cell |
| CD3^+^ CD8^+^ | CD8+ T cell (all) |
| CD3^+^ CD8^+^ CCR7^+^ CD45RA^–^ | Central memory CD8+ T cell |
| CD3^+^ CD8^+^ CCR7^–^ CD45RA^–^ | Effector memory CD8+ T cell |
| CD3^+^ CD8^+^ CCR7^+^ CD45RA^+^ | Naive CD8+ T cell |
| CD3^+^ CD8^+^ CCR7^–^ CD45RA^+^ | Effector CD8+ T cell |
| *B cell* |  |
| CD3^–^ CD19^+^ | B cell (all) |
| CD3^–^ CD19^+^ CD27^–^ IgD^–^ | Double negativ B cell |
| CD3^–^ CD19^+^ CD27^–^ IgD^+^ | Naive B cell |
| CD3^–^ CD19^+^ CD27^–^ IgD^+^ IgM^+^ | IgM^+^ naive B cell |
| CD3^––^ CD19^+^ CD21^low^ CD38^low^ | Exhausted B cells |
| CD3^–^ CD19^+^ CD27^+^ IgD^+^ | Non-switched memory B cell |
| CD3^–^ CD19^+^ CD27^+^ IgD^+^ IgM^+^ | IgM+ non-switched Memory B Cells |
| CD3^–^ CD19^+^ CD27^+^ IgD^–^ | Class-switched memory B cell (all) |
| CD3^–^ CD19^+^ CD27^+^ IgD^–^ IgA1^+^ | Class-switched memory B cell (IgA1) |
| CD3^–^ CD19^+^ CD27^+^ IgD^–^ IgA2^+^ | Class-switched memory B cell (IgA2) |
| CD3^–^ CD19^+^ CD27^+^ IgD^–^ IgG1^+^ | Class-switched memory B cell (IgG1) |
| CD3^–^ CD19^+^ CD27^+^ IgD^–^ IgG2^+^ | Class-switched memory B cell (IgG2) |
| CD3^–^ CD19^+^ CD27^+^ IgD^–^ IgG3^+^ | Class-switched memory B cell (IgG3) |
| CD3^–^ CD19^+^ CD38^high^ CD24^high^ IgM^high^ IgD^+^ | Transitional B cell |
| CD3^–^ CD19^+^ CD38^high^ CD24^high^ IgM^high^ IgD^+^  CD10^+^ | CD10+ transitional B Cells (T1 B Cells) |
| CD3^–^ CD19^+^ CD38^high^ IgM^–^ | Plasmablast |

### **Supplementary Table 2 -** Demographic and clinical characteristics of patients during the first and the second wave.

| **Variable** | **Early phase (n = 40)** | **Late phase (n = 22)** | ***p-value*** |
| --- | --- | --- | --- |
| Positive CAM-ICU, n (%) | 23 (56%) | 16 (73%) | 0.361 |
| Age in year, median (IQR) | 61 (57-69) | 63 (55-69) | 0.864 |
| Sex, female, n (%) | 9 (23%) | 8 (36%) | 0.383 |
| BMI, median (IQR) | 28 (25-31) | 29 (25-33) | 0.649 |
| SAPS II at admission, median (IQR) | 37 (31-43) | 37 (33-46) | 0.987 |
| Worst PaO_2_/FiO_2_, median (IQR) | 0.61 (0.53-0.77) | 0.58 (0.50-0.62) | 0.124 |
| Time of mechanical ventilation in days, median (IQR) | 13 (5-21) | 11 (8-25) | 0.889 |
| Total midazolam doses in mg/kg, median (IQR) | 0.92 (0-8.90) | 2.40 (0-10.61) | 0.652 |
| Total propofol doses in mg/kg, median (IQR) | 703 (192-1039) | 583 (334-1220) | 0.865 |
| Total fentanyl doses in ug/kg, median (IQR) | 232 (64-444) | 277 (138-518) | 0.581 |
| Received corticoids^a^ before sampling, n (%) | 1 (3%) | 22 (100%) | **< 0.001** |
| Received tocilizumab before sampling, n (%) | 31 (78%) | 5 (23%) | **< 0.001** |
| Received hydroxychloroquine before sampling, n (%) | 30 (75%) | 0 (0%) | **< 0.001** |
| Time between ICU admission and sampling, median (IQR) | 2 (1-5) | 3 (1-8) | 0.290 |

Abbreviation: CAM-ICU = Confusion Assessment Method for the Intensive Care Unit; IQR = interquartile; BMI = body mass index; SAPS II = Simplified Acute Physiology Score II; FiO_2_ = inspired oxygen fraction; PaO_2_ = oxygen partial pressure

^a^: Dexamethasone, prednisolone, or methylprednisolone

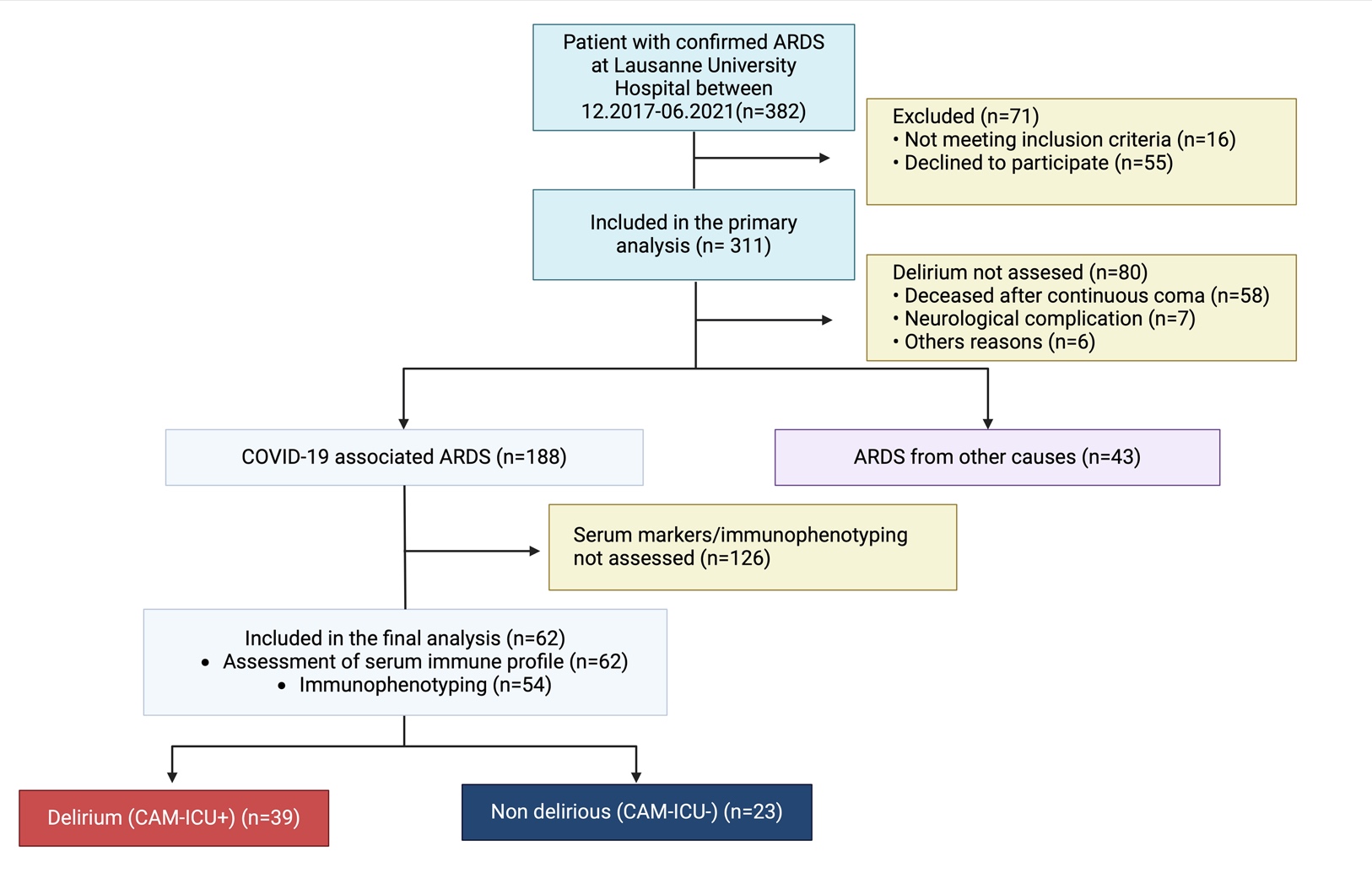

**Supplementary Figure 1 - Flow chart.** ARDS: acute respiratory distress. CAM-ICU: confusion assessment method in intensive care unit

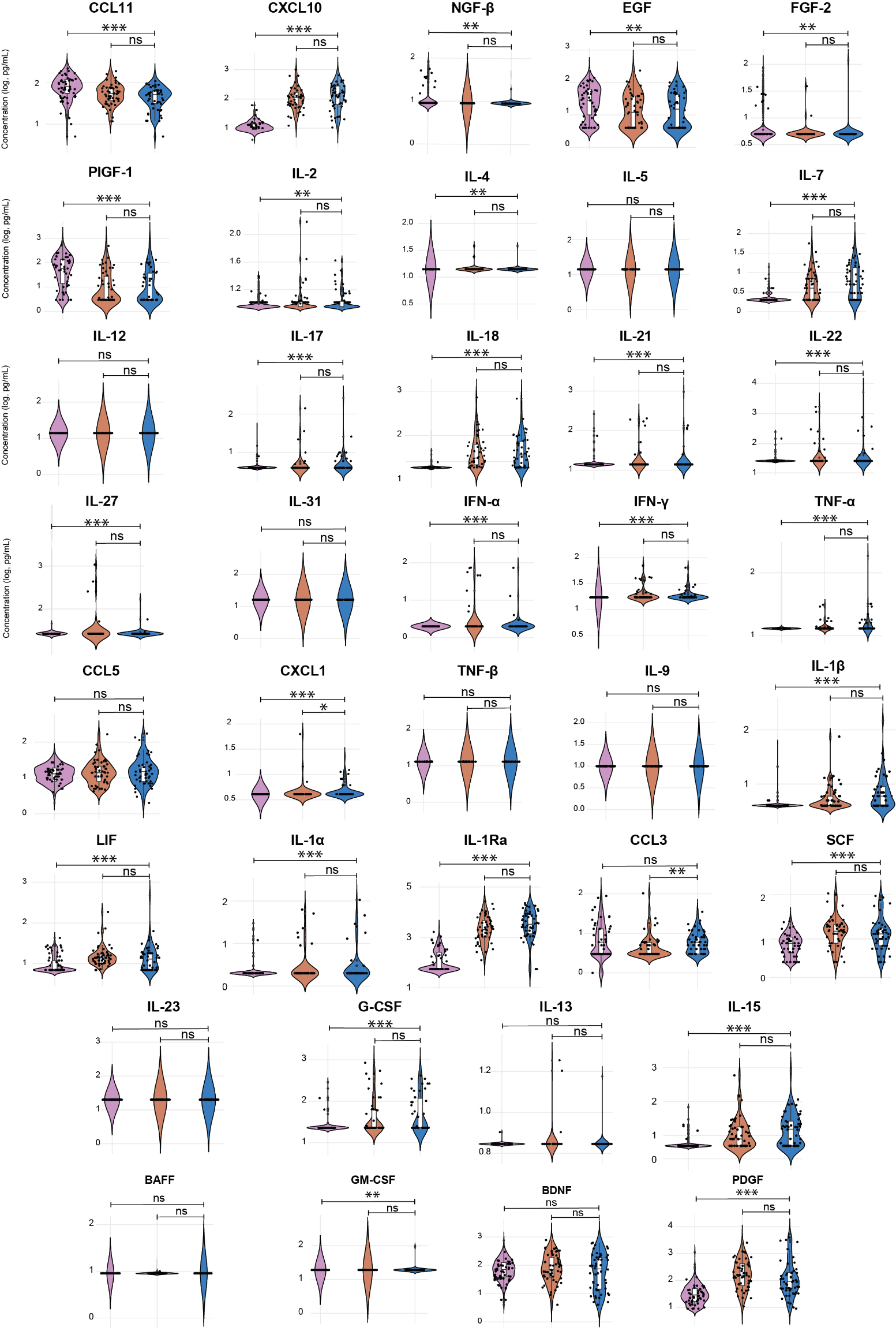

**Supplementary Figure 2 – Cytokines, Chemokines and Growth factors associated with COVID-19 and COVID-19 severity.**

Box and violin plots comparing concentrations of cytokines/chemokines/growth factors (after log10 transformation) in patients who developed severe-COVID-19 (blue, n=55) vs. non severe-COVID-19 (orange, n=62) and healthy controls (violet, n=450). Additionally, individual data points (n=50 per group) are displayed as scatter plots, randomly selected from each group. Statistical significance was calculated using the Mann-Whitney U test on the full dataset: *p < 0.05, **p < 0.01, ***p < 0.001, ns: not significant (p > 0.05). Soluble markers without variations across groups are not displayed.

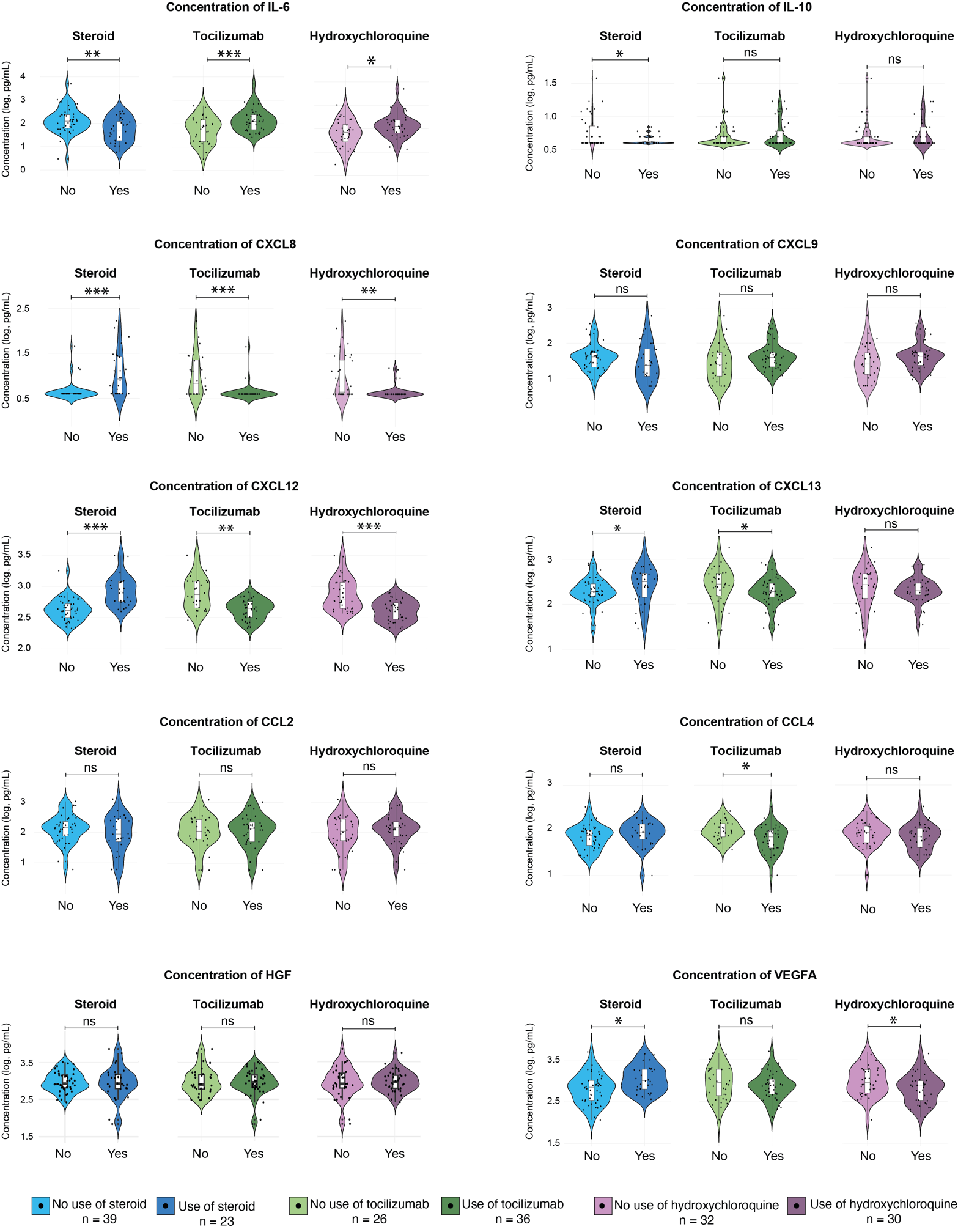

**Supplementary Figure 3 – Impacts of immunomodulatory treatments on serum soluble factors.**

Box and violin plots comparing concentrations of cytokines/chemokines/growth factors associated with COVID-19 severity (after log10 transformation) in patients that were treated with steroids (dark blue, n=23) or not (light blue, n=39), tocilizumab (dark green, n=36) or not (light green, n=26) and hydroxychloroquine (dark violet, n=30) or not (light violet, n=32). Statistical significance was calculated using the Mann-Whitney U test on the full dataset: *p < 0.05, **p < 0.01, ***p < 0.001, ns: not significant (p > 0.05).

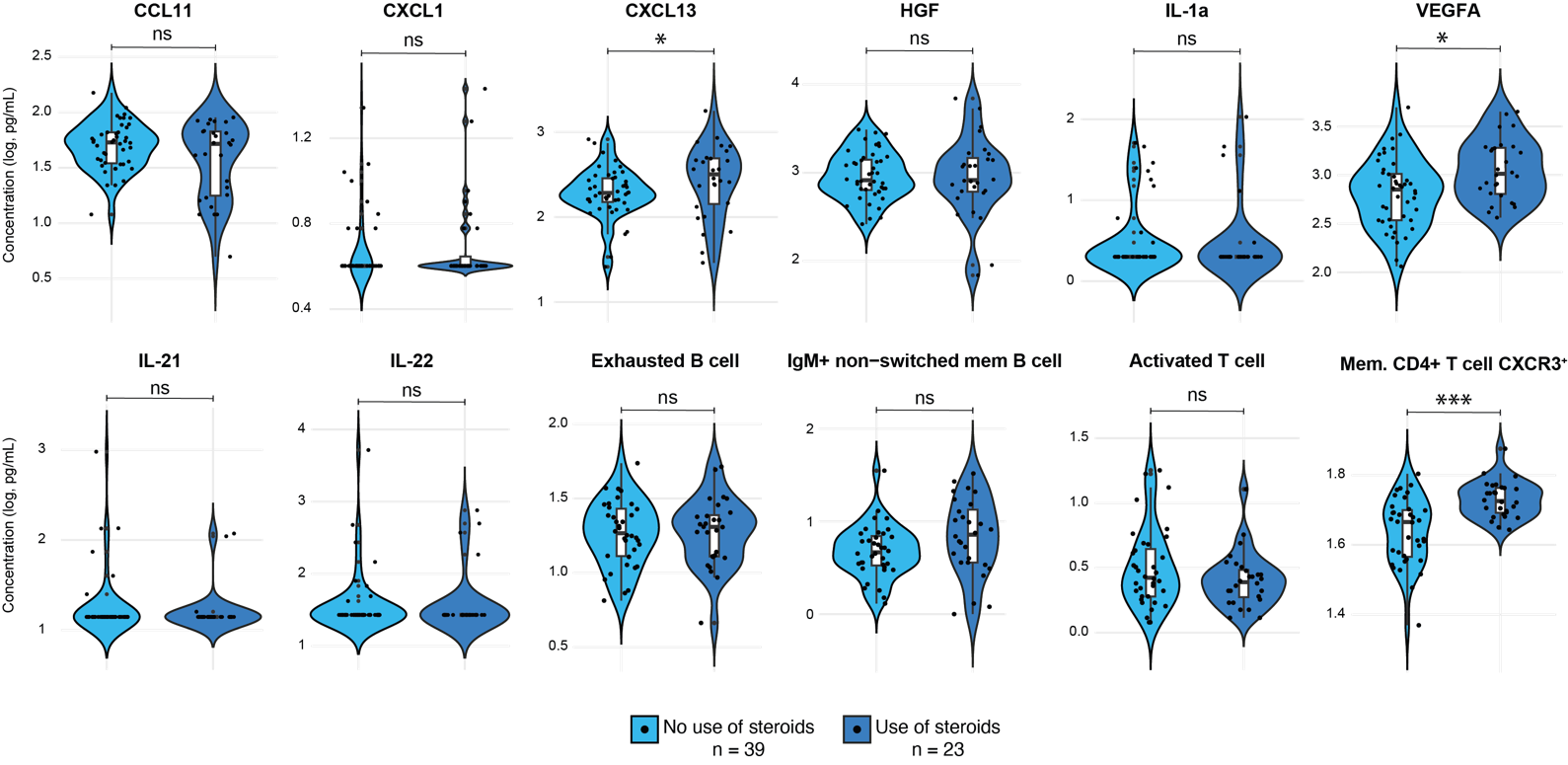

**Supplementary Figure 4 – Impact of immunomodulatory treatments on soluble factors and immune population associated with delirium.**

Box and violin plots comparing concentrations of cytokines/chemokines/growth factors and immune population associated with delirium in patients that were treated with steroids (dark blue, n=23) or not (light blue, n=39) prior sampling. Statistical significance was calculated using the Mann-Whitney U test on the full dataset: *p < 0.05, ***p < 0.001, ns: not significant (p > 0.05).
